# COVID-19 in Brazilian children and adolescents: findings from 21 hospitals

**DOI:** 10.1101/2021.08.08.21261510

**Authors:** Priscila Menezes Ferri Liu, Maria do Carmo Barros de Melo, Lilian Martins Oliveira Diniz, Cristiane dos Santos Dias, André Soares de Moura Costa, Bárbara Lopes Farace, Carla Thais Candida Alves da Silva, Fernando Anschau, Fernando Graça Aranha, Guilherme Fagundes Nascimento, Helena Duani, Jamille Hemétrio Salles Martins Costa, Karen Brasil Ruschel, Matheus Carvalho Alves Nogueira, Neimy Ramos de Oliveira, Roberta Pozza, Saionara Cristina Francisco, Thalita Martins Lage, Yuri Carlotto Ramires, Daniella Nunes Pereira, Zilma Silveira Nogueira Reis, José Miguel Chatkin, Milena Soriano Marcolino

## Abstract

**Introduction:** Children and adolescents with Covid-19 have been shown lower mortality less intense symptoms when compared to adults, but studies in Brazil have been based on the compulsory notifying system only.

**Objective:** To analyse clinical, laboratory, radiological characteristics and outcomes of hospitalized patients under 20 years with Covid-19.

**Methods:** Cases series of hospitalized patients with confirmed Covid-19 under 20 years, obtained from a cohort study in 37 hospitals from five states of Brazil.

**Results:** From 36 patients, 20 (55.5%) were adolescentes, 20 (55.5%) were male, 18 (50.0%) had comorbidities, 2 were pregnant and in 7 (19.4%), initial symptoms occurred during hospitalization for other causes, of whom 3 were possibly infected in the hospital. Fever (61.1%), dyspnea (33.3%) and neurological symptoms (33.0%) were the most common complaints. C-reactive protein was higher than 50mg/L in 16.7% and D-dimer was above the reference limit in 22.2%. Chest X-rays were performed in 20 (55.5%) patients, 9 had abnormalities, and chest tomography in 5. Hospital length of stay ranged from 1-40 days (median 5 [interquartile range 3-10]), 16 (44.4%) needed intensive therapy, 6 (16.7%) required mechanical ventilation and one patient (2.8%) died.

**Conclusion:** In case series patients under 20 years from hospitals from 5 states of Brazil, comorbidities were frequent, and most common symptoms were fever, dyspnea and neurological symptoms. Forty-four percent required intensive therapy, showing that the disease was not as mild as it was expected, and one patient died.

## I INTRODUCTION

The coronavirus disease 19 (COVID-19) pandemic has changed life and connections worldwide, leading to a global emergency.^1^ Disease spread has been hampered by social, cultural, financial and behavioural factors. In this context, some nations were more affected than others, and Brazil was one of the most severely hit countries.^1^

Overall, children have lower mortality and lower symptom intensity than adults, except for cases that progress to multisystem inflammatory syndrome in children (MIS-C). The MIS-C is an usual, but severe complication.^2^ It is associated with coagulopathies, macrophage activation syndrome and toxic shock syndrome, all of which contribute to increased severity and a higher risk of multiorgan failure, shock and mortality. Therefore, pediatricians should promptly diagnose and manage this condition.^3^

The first Covid-19 listed studies with pediatric patients were from China and the USA.^4,5^ Brazil is a continental country, with many socioeconomic disparities and cultural differences amongst its state’s populations. These contrasts were exacerbated by COVID-19, therefore, it is important to fully comprehend the disease’s demographic, clinical and laboratory characteristics, along with prognosis in hospitalized children and teenagers. An analysis using the compulsory notifying system SIVEP-Gripe of 67,180 confirmed COVID-19 patients from February to May 2020 identified that 2.2% of them were younger than twenty years.^3^ A study in Pernambuco state demonstrated high fatality rates in newborns and children under twelve months with severe conditions requiring hospitalization, in comparison to older infants.^6^ Another study using data from SIVEP-Gripe from February 2020 to January 2021 observed that, among 11,613 patients with age under twenty years, 7.6% died.^7^ However, all those studies were based one secondary data from the compulsory notifying system. Studies from other conditions have already shown that in practice, there is a flawed, incomplete filling. This may be due to impossibility of collecting the data, or by not understanding the reason for collecting that information, as it has not value for decision making.^8^

Therefore, data from cohort studies are important to provide more detailed characteristics of the clinical cases. Therefore, the objective of this study was to analyse clinical, laboratory and radiological characteristics, as well as patient outcomes, in patients with Covid-19 younger than twenty years who were hospitalized from a Brazilian cohort.

## II METHODS

This case series is a substudy of the Brazilian Covid-19 Registry, a multicentric cohort study^9^, which included 38 hospitals, 37 of them were active at the time of assessment. In 21 of them, children and teenagers younger than twenty years were treated. From these, eight were public and thirteen were private hospitals. The 21 hospitals were from six municipalities in the states of Minas Gerais (cities of Belo Horizonte, Curvelo and Ipatinga), Rio Grande do Sul (Porto Alegre and Bento Gonçalves) and Santa Catarina (Florianópolis).

The study has been conducted according to a predefined protocol and was approved by the National Commission for Research Ethics (CAAE 30350820.5.1001.0008).^8^ Due to the state of pandemic and to the fact that data was solely collected through medical records, based on unidentified data, individual informed consent was waived.

This manuscript adheres to the STROBE guidelines (*Strengthening the Reporting of Observational Studies in Epidemiology*).^10^

### Data collection

Consecutive patients with a confirmed diagnosis by RT-PCR or by positive serological tests (IgM) for SARS-CoV-2, according to the World Health Organization (WHO)^11^ criteria and who were admitted to participating hospitals from March to September 2020 were enrolled. Trained healthcare professionals or medical and nursing undergraduate students collected data using Research Data Capture (REDCp) tools.^12^

Medical records were assessed by trained and supervisioned hospital staff or undergraduate students in order to collect data regarding demographics and clinical characteristics, comorbidities, medications in use, symptoms and clinical evaluation upon hospital admission, laboratory and radiological exams, medication used throughout hospitalization and outcomes.

The primary outcome was in-hospital mortality. Secondary outcomes included time spent in the Intensive Care Unit (ICU), time spent in the hospital, clinical complications and the need for additional life support resources.

### Statistical analysis

The Shapiro-Wilk normality test was performed to determine whether the continuous variables were normally distributed. As all variables had non-normal distribution, they were summarized using medians and interquartile ranges (IQR). Categorical variables were summarized as counts and percentages. Statistical analyses were conducted IBM SPSS Statistics (IBM SPSS Statistics for Macintosh, Version 19 Armonk, NY: IBM Corp.).

## III RESULTS

From the 7,346 Covid-19 hospital registrations on the database, 36 patients were under twenty years (0.5%), who were included in the analysis. Of those, 20 were adolescentes (55.5%), 20 were males (55.5%) and two of the females were pregnant. One of them was 35 weeks and the other 36 weeks pregnant. All of them had Covid-19 diagnosis confirmed by RT-PCR.

Eight patients (22.2%) were admitted through transfer from other hospitals, three (8.3%) from emergency care units, one (2.8%) from other institutions on the same service, four (11.1%) from an institution from another municipality and the others sought care straight in the participating hospital. With regards to patient origin, Belo Horizonte was the most common one (10/27.8%), followed by Porto Alegre (3/8.3%), Ipatinga (3/8.3%) and Bento Gonçalves (3/8.3%). Data on demographics and clinical characteristics are shown in Table 1.

**Table 1.** Data related to age, sex, commorbidities, lifestyle habits, registration of surgical procedure and use of continuous medication in analysed patients

| Variables | Total (N=36)<br>N (%) |
| --- | --- |
| **Age median (IQ25%- IQ75%)** | 14 years<br>(11 months – 16 years) |
| **Male** | 16 (44.5%) |
| **Comorbidities** | (N=36) |
| Cerebral Paralysis | 4 (11.1%) |
| Asthma | 3 (8.3%) |
| Epilepsy | 3 (8.3%) |
| Diabetes Mellitus | 2 (5.5%) |
| Obesity | 1 (2.8%) |
| Psychiatric Disease | 1 (2.8%) |
| HIV Infection | 1 (2.8%) |
| Hematológica Malignant Neoplasm | 1 (2.8%) |
| *Other relevant health condition | 2 (5.5%) |
| **Lifestyle Habits** |  |
| Ex-Smoker | 2 (5.5%) |
| Alcoholism | 0 |
| Use of Illicit Drugs | 0 |
| **Surgical Procedures** |  |
| On the last 90 days | 1 (2.8%) |
| **Continuously Used Drugs In Domicile** |  |
| Anticonvulsant | 6 (16.7%) |
| Long-acting Bronchodilator | 2 (5.5%) |
| Inhalatory Corticoid | 1 (2.8%) |
| Oral Hypoglycemiant | 1 (2.8%) |
| Immunosuppressant | 1 (2.8%) |
| Insulin | 1 (2.8%) |
| **Others | 5 (13.9%) |

\*Other relevant health condition: anemia (N=1); wheezing infant (N=1)
\*\* Other continuously used drugs: anxiolytic (N=2); trimethoprim sulfamethoxazole (N=1); antidepressant (N=1); desmopressin (N=1)

In seven (19,4%) cases, the initial symptoms of Covid-19 began during hospitalization for other reasons, and three of these (8.3% of the total) were infected while in the hospital. Reasons for hospitalization were diverse and concerned premature childbirth, with newborns asymptomatic for Covid-19 (n=1), abdominal pain (n=1), premature rupture of membranes and childbirth with newborn’s death due to birth complications (n=1), elective surgery (n=1) and convulsive crisis (n=3). Table 2 displays clinical data of patients upon hospital admission. The most common symptoms were: fever, dyspnea, headache, rhinorrhea, dry cough and neurological symptoms.

**Table 2.** Data present in the initial clinical evaluation of the analysed population

| Variables | Total (N=36)<br>N (%) |
| --- | --- |
| **Symptoms** |  |
| Fever | 22 (61.1%) |
| Dyspnea | 12 (33.3%) |
| Neurological symptoms | 7 (19.4%) |
| Dry cough | 7 (19.4%) |
| Rhinorrhea | 7 (19.4%) |
| Hyporexia | 6 (16.7%) |
| Diarrhea | 6 (16.7%) |
| Adynamia | 6 (16.7%) |
| Nausea/vomiting | 5 (13.9%) |
| Headache | 5 (13.9%) |
| Productive cough | 4 (11.1%) |
| Irritability | 4 (11.1%) |
| Sore throat | 2 (5.5%) |
| Ageusia | 1 (2.8%) |
| Anosmia | 1 (2.8%) |
| Myalgia | 1 (2.8%) |
| *Others | 7 (19.4%) |
| **Clinical data at hospital presentation** |  |
| Glasgow <15 | 7 (19.4%) |
| Peripheral oxygen saturation<br>≤94% | 6 (16.7%) |
| Tachypnea (age referenced) | 5 (13.9%) |
| Fever | 5 (13.9%) |
| Tachycardia (age referenced) | 3 (8.3%) |
| Supplemental oxygenation |  |
| None | 26 (72.2%) |
| Nasal cannula 1-4 L/min | 7 (19.4%) |
| Facial mask (5-15 L/min) | 2 (5.5%) |
| Mechanical ventilation | 1 (2.8%) |

\*Other symptoms: convulsive crisis (N=1); hypoactivity in newborns (N=2); abdominal pain/ abdominal distension (N=2); purulent discharge in tracheostomy (N=1); sneezes (N=1); chest pain (N=1).

At hospital admission, chest radiography was performed on twenty patients (55.5%). Of those, nine (25.0%) of them had abnormalities. The following abnormalities were observed: focal interstitial infiltration (n=4, 11.1%); parenchymal consolidation (n=2, 5.5%); bilateral ground-glass opacity (n=2, 5.5%); and interstitial diffuse infiltration (n=1, 2.8%). In five patients (13.9%), a chest tomography was requested. The most common abnormalities were athelectasy (n=1, 2.8%); lung parenchymal consolidation (n=1, 2.8%); pleural effusion (n=1, 2.8%); unilateral ground-glass opacity (n=1, 2.8%); bilateral ground-glass opacity (n=1, 2.8%); and peripheral ground-glass opacity (n=1, 2.8%). C-reactive protein was above 50mg/L in 16.7% and D-dimer was above the reference limit in 22.2%. Results of other exams are shown in Table 3.

**Table 3.** Laboratory parameters of the studied population at the hospital’s admission time considering medium values of reference that comply with the age range evaluated

| Variables | Median<br>(P25-P75) | Mean<br>(SD) | Minimum -<br>Maximum |
| --- | --- | --- | --- |
| Hemoglobin (g/dL) | 12.25 (10.65 – 13.78) | 12.3 (±2.46) | 6.3 – 16.6 |
| Leukocyte count<br>(cels/mm<sup>3</sup>) | 8,320 (6,540 - 10,530) | 9,212 (±4,871) | 2,800 - 23,100 |
| Neutrophils (cels/mm<sup>3</sup>) | 5,789 (2,759 – 7,814) | 5,908 (±3,839) | 700 – 17,355 |
| Lymphocytes (cels/mm<sup>3</sup>) | 1,642 (1,179 – 3,133) | 2,106 (±1,364) | 343 - 6,900 |
| Platelets (x10<sup>3</sup>/L) | 236.50 (158.75 – 302.50) | 235.82 (±940.39) | 230 - 431 |
| Creatinine (mg/dL) | 0.58 (0.46 – 0.98) | 0.82 (±0.58) | 0.23 – 2.37 |
| Urea (mg/dL) | 23 (12 – 30) | 22 (±10.94) | 5 – 49 |
| Lactate (mmol/L) | 1.46 (1.22 – 1.95) | 1.93 (±1.47) | 0.60 – 5.14 |
| Lactic dehydrogenase | 300 (168 – 355) | 267.2 (±126.7) | 17 – 490 |
| C-reactive protein (mg/L)** | 22.30 (5.34 – 63.45) | 38.21 (±51.41) | 0.71 – 257 |
| pH | 7.41 (7.37 – 7.46) | 7.42 (±0.08) | 7.28 – 7.61 |
| pCO<sub>2</sub> | 32.50 (26.43 – 37.25) | 31.56 (±8.60) | 15.70 – 47.00 |
| pO<sub>2</sub> | 100.3 (77.35 – 161) | 112.7 (±44.6) | 42.0 – 194.0 |
| Bicarbonate (mEq/L) | 20.55 (15.15 – 23.38) | 19.96 (±4.90) | 12.6- 29.0 |

\* There was no request for procalcitonin and only on patient had ferritin ad admission (134.1 mcg/dL)
\*\* Six patients had C-reactive protein > 50 mg/L(16.7%).

The time from symptom onset to hospital presentation ranged from one to ten days, with a median of two days (IQR 1-4). Hospital length of stay ranged from one to forty days, with a median of five (interquartile range [IQR] 3-10) days. Table 4 summarizes medications and supportive care during hospital stay. During hospitalization, one patient died (2.8%). This patient was being treated for hematological cancer, using imunossupressor agent, and developed his first Covid-19 symptoms while in hospital to investigate abdominal pain. His condition deteriorated due to septic shock, and he died.

**Table 4.**
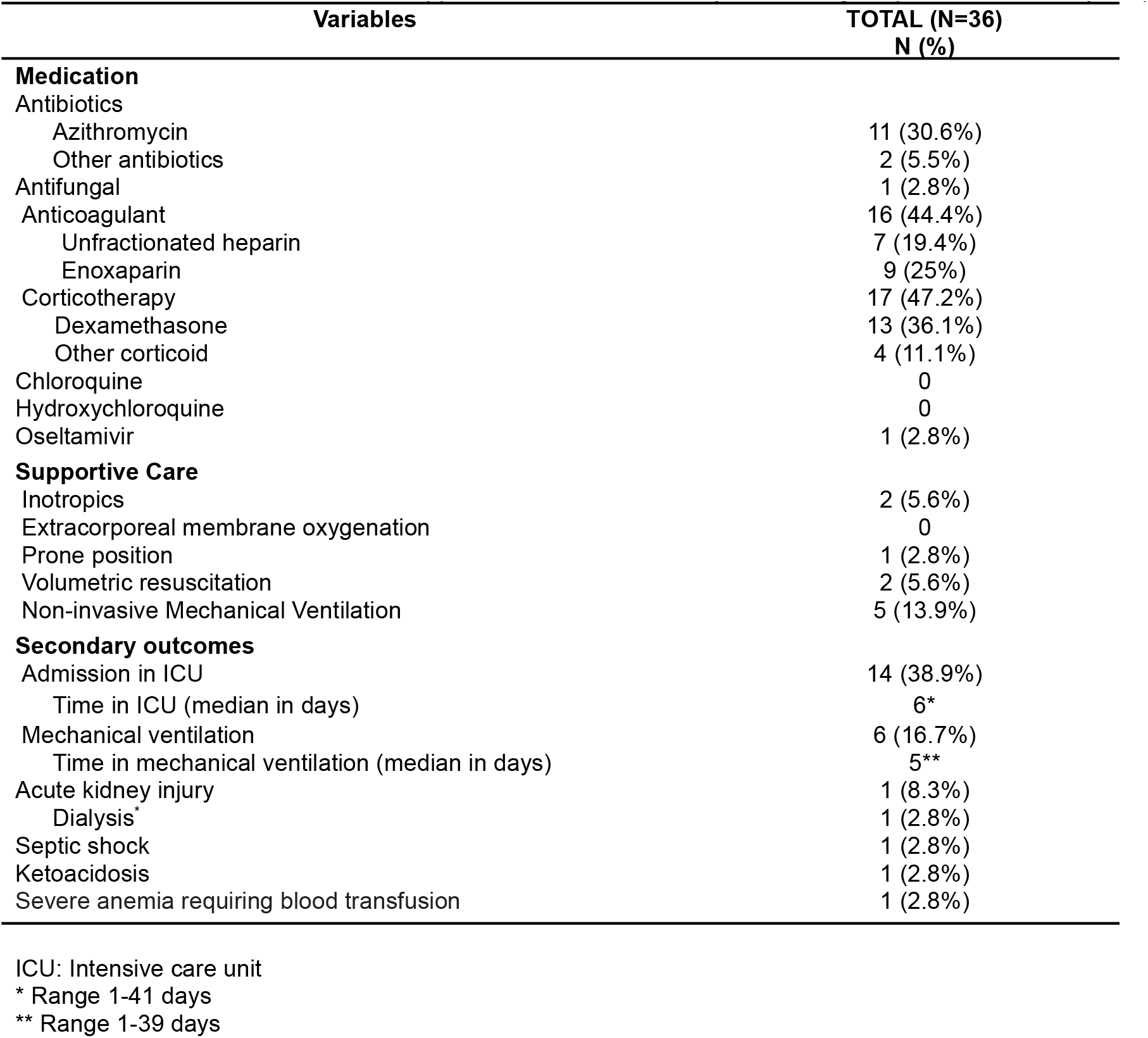
Administered medication, supportive care and secondary data during hospitalization of analysed patients

| Variables | TOTAL (N=36)<br>N (%) |
| --- | --- |
| **Medication** |  |
| Antibiotics |  |
| Azithromycin | 11 (30.6%) |
| Other antibiotics | 2 (5.5%) |
| Antifungal | 1 (2.8%) |
| Anticoagulant | 16 (44.4%) |
| Unfractionated heparin | 7 (19.4%) |
| Enoxaparin | 9 (25%) |
| Corticotherapy | 17 (47.2%) |
| Dexamethasone | 13 (36.1%) |
| Other corticoid | 4 (11.1%) |
| Chloroquine | 0 |
| Hydroxychloroquine | 0 |
| Oseltamivir | 1 (2.8%) |
| **Supportive Care** |  |
| Inotropics | 2 (5.6%) |
| Extracorporeal membrane oxygenation | 0 |
| Prone position | 1 (2.8%) |
| Volumetric resuscitation | 2 (5.6%) |
| Non-invasive Mechanical Ventilation | 5 (13.9%) |
| **Secondary outcomes** |  |
| Admission in ICU | 14 (38.9%) |
| Time in ICU (median in days) | 6* |
| Mechanical ventilation | 6 (16.7%) |
| Time in mechanical ventilation (median in days) | 5** |
| Acute kidney injury | 1 (8.3%) |
| Dialysis | 1 (2.8%) |
| Septic shock | 1 (2.8%) |
| Ketoacidosis | 1 (2.8%) |
| Severe anemia requiring blood transfusion | 1 (2.8%) |

ICU: Intensive care unit
\* Range 1-41 days
\*\* Range 1-39 days

## IV DISCUSSION

Despite previous publications in large databases of children and adolescents with Covid-19 in Brazil, this series provided more detailed data on clinical characteristics, laboratory and imaging data, as well as hospital outcomes.

It was observed that cases in patients younger than 20 years were rare in this hospital cohort (0.5%). According to a recent systematic review of studies in pediatric patients, 21 to 25% of all cases were from six to fourteen years-olds.^13^In the present study, half of hospitalized patients were older than 14 years old, but 25% were under eleven months old, showing the importance of the age group with less than a year old in hospitalizations.

It is still unexplained and intriguing why children and adolescents are more frequently asymptomatic or had less severe clinical forms of Covid-19. There are some hypothesis to explain this findings, including less expression of virus receptors, recent exposure to other viruses and cross protection, more developed innate immunity, among other factors.^14^ However, it is important to highlight the evidence that susceptibility to infection is similar in all age groups, so children can disseminate the virus in the community even when asymptomatic.^13^

In the current study, fever was the predominant symptom in children hospitalized with Covid-19 (61.1%), followed by dyspnea (33.3%). These findings, as well as cough, were less frequent than what was observed in the large sample of patients in the aforementioned study based on data collected from the compulsory form.^7^ We hypothesized that, in those studies, physicians tended to notify in the compulsory system patients who were more symptomatic. Our findings were in line with the European cohort, in which fever was observed in 65% of patients and symptoms of superior respiratory tract infection in 54%. Only 25% of children had symptoms of a lower respiratory tract infection, 28% had headaches and 22% had gastrointestinal symptoms.^15^ Diarrhea, nausea, vomiting and anorexia have been reported in up to 15% of Covid-19 patients, predominantly in children under the age of three.^13,14,16^

It was also observed in this study that 33% of children experienced neurological symptoms while in the hospital. Several studies in adults and in children have linked Covid-19 to a variety of central and peripheral neurological insults, ranging in severity from a mild headache and/or anosmia to peripheral neuropathy, seizures, stroke, demyelinating disorders and encephalopathy.^17^ The long-term impact of neurological damage in those patients who develop neurological symptoms remains uncertain.^17^

Comorbidities have been associated with the development of more severe cases of the disease. Half of our patients had comorbidities, which is more than what were reported in the study based on the SIVEP-Gripe database (28.1%).^7^ Cerebral paralysis (11.1%), epilepsy (8.3%) and asthma (8.3%) were the most common comorbidities in the present analysis, while in the SIVEP-Gripe study, asthma (7.5%) and neurological disorders (6.1%) were also the most common ones.

Other studies showed that patients with comorbidities had a 1.79 times greater risk of being admitted to intensive care units and a 2.8 times greater risk of dying.^18,19^ Obesity, chronic pulmonary disease, cardiovascular disease, neurological disease and immunological disease were all associated to worse disease progression in children in various studies.^18,19^ Obesity has been shown to induce high levels of local tissue and systemic inflammation, as well as elevating cytosine levels related to SARS-CoV-2 infection pathogenesis.^18^ Since only one patient in the current study was diagnosed as obese, this risk relationship could not be established.

The two pregnant adolescents with Covid-19 who gave birth to three newborns, two of them were asymptomatic to Covid-19 and the other died due to preterm birth complications. The majority of women with Covid-19 are asymptomatic and, if they are pregnant, the impact on the fetus and the newborn is still unknown.^20^ Symptomatic pregnant women, on the contrary, when compared to those without Covid-19 with the same gestational age, have 22 times higher risk of death.^21^In Brazil, the rate of maternal mortality due to Covid-19 is 13.4% in 2021, with a relevant increase in comparison to 2020 (6.4%).^22^ There is also a higher risk of preeclampsia, especially among nulliparous women, independent of any other risk factors or preexisting conditions, independent of Covid-19’s severity.^21^ Preeclampsia and Covid-19 have shown to be independently and additively associated with premature childbirth, severe perinatal morbidity and mortality and adverse maternal outcomes.^23^In addition, Covid-19 increases the risk of fetal distress, admission to intensive care unit (ICU) and severe newborn complications.^21^Infected pregnant women, especially the ones who develop pneumonia, also seem to have an increased premature childbirth frequency and possibly require a cesarean. These complications are probably related to severe maternal diseases^24^, however, it is still under investigation whether there is intrauterine infection. Placental infections and newborn infections have been reported, however the possibility of intrauterine vertical transmission of SARS-Cov-2 is still controversial.^25,26^

Previous studies, including infections caused by other coronaviruses, have reported normal or reduced leukocyte values, elevated lactate dehydrogenase (LDH) and D-dimer. The study by Balad *et al*. (2020)^13^ also showed a high prevalence of leukopenia and lymphopenia. In that study, the most common abnormal markers were elevated procalcitonin and ferritin, consistent with an exacerbated inflammatory state. These markers, however, are not specific, limiting their clinical utility in the absence of clinical and epidemiological factors that suggest infection.^13^In the present study, the request for laboratorial exams were at the discretion of attending physicians, so procalcitonin and ferritin was not requested, but 16.8% had shown C-reactive protein and 22.2% had elevated D-dimer. There were no significant abnormalities in the leukocytes and lymphocytes counts, even for the patient who died.

With regards to radiological findings, in the European study from Gotzinger *et al*. (2020), 47% of children that had a chest radiograph presented abnormalities that suggested pneumonia.^15^ In the present study, we observed that 45% of patients presented chest X-ray abnormalities, in line with other studies. The radiological findings have been highly inespecific and provide little evidence regarding individual SARS-CoV-2 infection, not allowing to diagnose or to discard SARS-CoV-2 infection based in any of these findings.^13^

Most studies indicate good outcomes after infection in children, with rare fatalities, according to the typically mild cases observed in this age group. We observed that 38.9% of the study sample were admitted to ICU. In the aforementioned European study, only 8.2% required intensive care treatment.^12^ We believe that the high rates of ICU admissions observed in our study were related to concerns about the disease’s evolution in the pediatric age group and not to the severity, since the study shows that only 16.7% used invasive mechanical ventilation and 5.5% required the use of inotropic. Additionally, the two pregnant teenagers and two newborns included in the sample were also admitted to improve monitorization. Admission to intensive care has been more frequently described in younger children such as the newborns.^27^ Gotzinger *et al*.(2020)^15^ observed an association between age below a month old, male gender, lower respiratory tract symptoms and preexisting conditions with the need for ICU admission.

Except for rare cases of severe disease in patients with comorbidities and cases of MIS-C, the majority of patients have a good outcome.^15,26^ A meta-analysis revealed that in only 5% of cases children required hospitalization, with an estimated mortality of 0.3%.^13^In 2020, children accounted for 2.5% hospitalizations and 0.6% of total deaths by Covid-19 in the country.^14^In the aforementioned large study based on data from SIVEP-Gripe from February 2020 to January 2021, mortality was 7.6% By multivariate analysis, age less than 2 years or adolescents (12 to 19 years old) had higher risk of death. In the age group of 2 to 11 years old, most deaths were from indigenous ethnicity, residents of Northeast or North regions (underprivileged areas) of the country when compared to deaths from the Southeast region; and patients with pre-existing diseases.^7^

The present study has limitations, as it is a case series, based on a multicenter retrospective cohort. However, it adds knowledge to symptoms, clinical presentation, laboratory results and imaging findings, as well as patient outcomes in this population. New studies are needed for a better understanding of disease evolution in the pediatric population, as well as in pregnant women and newborns.

In conclusion, this case series of Covid-19 patients under 20 years from hospitals from 5 states of Brazil, comorbidities were frequent, and most common symptoms were fever, dyspnea and neurological symptoms. Forty-four percent required intensive therapy, showing that the disease was not as mild as it was expected, and one patient died. Further studies are required to further investigate aspects of the disease in this population.

## Data Availability

Data is available through reasonable solicitation.
Access, responsibility and data analysis
The main authors (MSM and MCBM) had access to the entire study 's data and took on responsibility for data integrity and precision in data analysis.

## DECLARATIONS

### Ethics approval and participation consent

The study protocol was approved by the National Commission on Research Ethics (CAAE 30350820.5.1001.0008). Individual consent was dismissed due to the severity of the situation and because of the use of unidentified data, based only on the revision of medical records.

### Declaration of data availability

Data is available through reasonable solicitation.

### Access, responsibility and data analysis

The main authors (MSM and MCBM) had access to the entire study’s data and took on responsibility for data integrity and precision in data analysis.

### Declaration of transparency

The main authors (MSM and MCBM) claim that the manuscript is an honest repost, precise and transparent of the related study; that no important aspect of the study was omitted; and that any discrepancies regarding the originally planned study (and, if relevant, registered) were explained.

## AUTHOR CONTRIBUTIONS

Substantial contributions for the conception or design of the manuscript: MSM, MCBM, PMFL, LD and CSD.

Substantial contributions for data acquisition, analysis or interpretation: MSM, MCBM, PMFL, LD, CSD, ASMC, BLF, CTCAS, FA, FGA, GFN, HD, JHSMC, JMC, KBR, MCAN, NRO, RP, SCF, TML, YCR.

Manuscript formulation: MSM, MCBM, PMFL, LD, CSD and DNP.

Critically revised the manuscript in relation to the important intellectual content: all authors. Final version’s approval: all authors

Agreed to be responsible for all aspects of the work, ensuring that issues related to precision of integrity in any of the work’s parts will be properly investigated and solved: MSM and MCBM.

## ACKNOWLEDGEMENTS

We acknowledge the hospitals that take part in the research project National Multicentric Hospital Registry of Patients with diseases caused by SARS-COV-2 (COVID 19), that originated this work: Hospital Bruno Born; Hospital das Clínicas da UFMG; Hospital de Clínicas de Porto Alegre; Hospital Santo Antônio; Hospital Eduardo de Menezes; Hospital Mãe de Deus; Hospital Márcio Cunha; Hospital Mater Dei Betim-Contagem; Hospital Mater Dei Contorno; Hospital Mater Dei Santo Agostinho; Hospital Metropolitano Dr. Célio de Castro; Hospital Metropolitano Odilon Behrens; Hospital Moinhos de Vento; Hospital Nossa Senhora da Conceição; Hospital Risoleta Tolentino Neves; Hospital São Lucas da PUCRS; Hospital Sempre; Hospital SOS Cárdio; Hospital Tacchini; Hospital Unimed-BH; Hospital Universitário de Santa Maria.

We also acknowledge the clinical staff of these hospitals, that cared for the patients and to all the undergraduate students that aided in data collection.

